# Fast and accurate diagnostics from highly multiplexed sequencing assays

**DOI:** 10.1101/2020.05.13.20100131

**Authors:** A. Sina Booeshaghi, Nathan B. Lubock, Aaron R. Cooper, Scott W. Simpkins, Joshua S. Bloom, Jase Gehring, Laura Luebbert, Sri Kosuri, Lior Pachter

**Affiliations:** Department of Mechanical Engineering, California Institute of Technology, Pasadena, CA; Octant Inc., Emeryville, CA; Department of Human Genetics, University of California, Los Angeles, Los Angeles, United States; Department of Genome Sciences, University of Washington, Seattle, WA; Division of Biology and Biological Engineering, California Institute of Technology, Pasadena, CA; Department of Computing and Mathematical Sciences, California Institute of Technology, Pasadena, CA

## Abstract

Scalable, inexpensive, accurate, and secure testing for SARS-CoV-2 infection is crucial for control of the novel coronavirus pandemic. Recently developed highly multiplexed sequencing assays that rely on high-throughput sequencing (HMSAs) can, in principle, meet these demands, and present promising alternatives to currently used RT-qPCR-based tests. However, the analysis and interpretation of HMSAs requires overcoming several computational and statistical challenges. Using recently acquired experimental data, we present and validate an accurate and fast computational testing workflow based on kallisto and bustools, that utilize robust statistical methods and fast, memory efficient algorithms for processing high-throughput sequencing data. We show that our workflow is effective at processing data from all recently proposed SARS-CoV-2 sequencing based diagnostic tests, and is generally applicable to any diagnostic HMSAs.

## Introduction

Rapid, scalable, low-cost testing for SARS-CoV-2 is paramount for reducing infection rates and controlling the current pandemic^1^. Currently, SARS-CoV-2 tests are primarily based on RT-qPCR, however, several groups have recently proposed massively parallelized diagnostic assays based on high-throughput sequencing that hold the promise of greatly increased throughput, reduced cost, and improved sensitivity^2–4^. While the proposed diagnostics differ in implementation details, they share several key features:

- (Synthetic) sequence barcodes known as sample indices are associated with samples and are recovered by sequencing.
- (Biological) sequences associated with genes, including viral genes, control genes, or spike-ins, are recovered by sequencing.
- Sequenced sample indices and biological sequences are associated with each other.

These assays bear resemblance to multiplexed barcoding technologies used for single-cell RNA-seq^5–7^, and as a result, the bioinformatics challenges that must be overcome in analyzing the data are similar.

Processing of the data requires association of the biological sequences with their genes of origin, error correction of the samples indices and collation of sequences associated with a single sample to count the number of molecules from each gene that have been observed (**Figure 1**). Finally, the infection status for each sample must be determined from the gene abundance estimates per sample.

**Fig 1:**
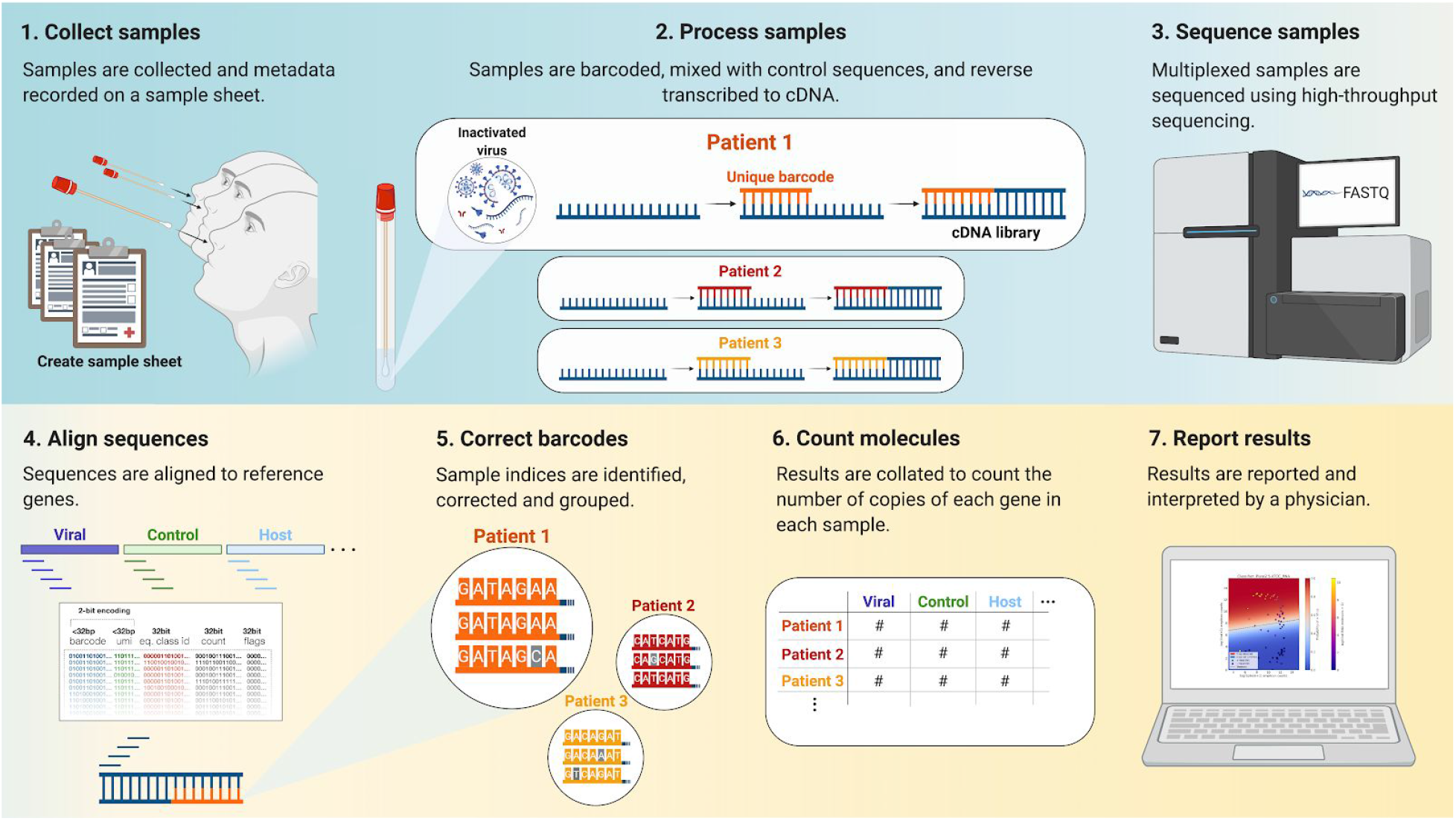
Massively parallel diagnostic testing by high-throughput sequencing. Workflow of a high-throughput sequencing based diagnostic test. 1) Samples are collected and prepared. 2) Samples are barcoded and amplified. 3) Multiplexed samples are pooled and sequenced using a high-throughput sequencer. 4) Sequencing data is aligned to a set of genes, 5) sample indices are error corrected, 6) counts are computed, and 7) diagnostic results are obtained.

To overcome these challenges, we adapted the RNA-seq and single-cell RNA-seq tools kallisto^8^ and bustools^9,10^ to HMSA analysis, and coupled them in a workflow we designate “kallisto | bustools” (**Supplementary Note**). In addition, we developed a testing framework to report infection status, and we validate our results with complementary methods. Our software is freely available under the permissible BSD-2 open source license, and we show that it can be used for SwabSeq^2^, a technology based on Octant’s RNA amplicon sequencing platform, LAMP-seq^3^, which relies on LAMP^11^, covE-seq ^12^, which targets the SARS-CoV-2 E gene, or TRB-seq^4^, which is a targeted BRB-seq^13^ variant. The short running time and low memory footprint of the software allow low-cost logistical solutions to data analysis that are important in the clinical setting.

## Results

To validate our workflow, we analyzed 307,494,992 SwabSeq reads (see Methods). This dataset consisted of two 384-well plates each with a titration series of viral RNA from two companies, Twist and ATCC, for a total of 768 uniquely barcoded samples. HEK293 lysate, nasopharyngeal (NP) lysate, and controls were included in all of the wells of each plate. The first plate was used to test primers bound to the SARS-CoV-2 N gene and the second plate was used to test primers bound to the SARS-CoV-2 S gene (**Supplementary Figure 1**). Reads were aligned to a custom set of reference sequences (see Methods) using kallisto, and sample indices were corrected to a whitelist. Finally, counts of genes per sample were collated to make a sample by gene matrix (see Methods) and this was used to determine, for each sample, whether it contained viral RNA.

**Figure 2a** shows the predicted classification results for the Plate 2 S ATCC RNA experiment obtained by training a logistic regression classifier on half of the data and testing on the remaining half. The classifier learns coefficients for each covariate that optimally (by the logistic model) classify positive versus negative samples (see Methods). Crucially, the model provides a probability for each classification. Furthermore, the weights estimated in the logistic regression make an intuitive visualization of standard curves possible where virus and spike-in, suitably normalized according to regression coefficients, are measured relative to one another (**Figure 2b**). This allows the assessment of the quality of a diagnostic assay in the context of classification via a standard curve.

**Figure 2:**
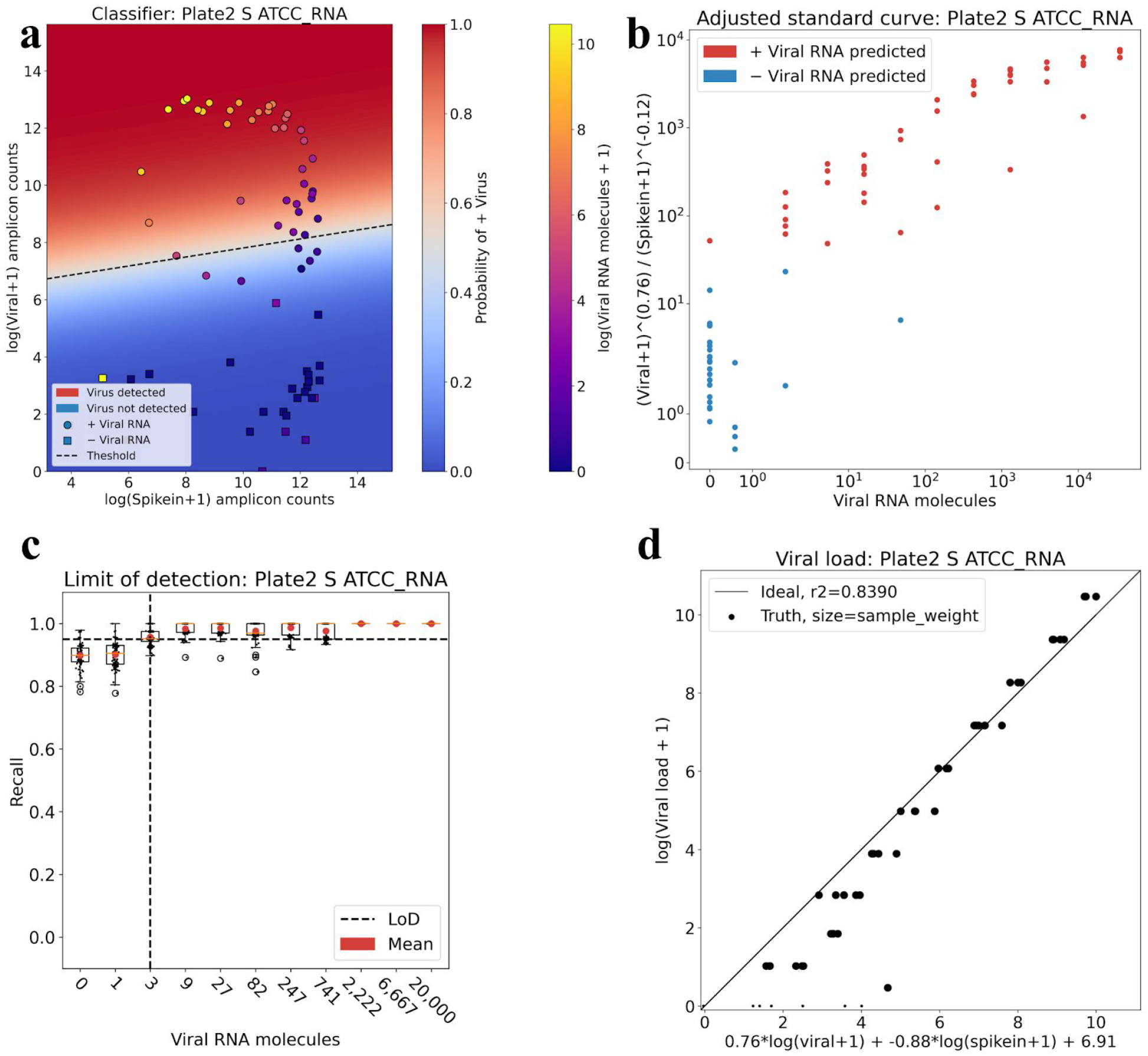
Sample classification, viral load prediction and limit of detection. **a)** Positive and negative samples from the Plate 2 S ATCC RNA experiment can be effectively separated using logistic regression. Points correspond to samples and are colored by the known amount of viral RNA per sample. The probability of each sample of having a non-zero amount of viral RNA is given by the logistic function and is painted as orthogonal to the logistic regression boundary. The shape of the point indicates whether it was predicted to be positive for viral RNA (circle) or negative (square). **b)** The standard curve measuring spike-in and virus vs the known amount of viral RNA per sample with optimal exponential coefficients determined by logistic regression; samples are colored by their predicted classification. **c)** The limit of detection as estimated from 99 rounds of split/test and logistic regression to classify samples with non-zero amount of viral RNA. The limit of detection is defined as the number of RNA molecules for which the recall is greater than 19/20 (=0.95) **d)** The viral load per sample can be predicted with (a weighted) linear regression using the log counts from each gene. Each point is a sample, with perfect predictions lying on the diagonal line. Size of points represents their weight, with points weighted so that each titer is represented with equal weight. The code to reproduce each figure is here: [code (a) and code (b)](https://github.com/pachterlab/BLCSBGLKP_2020/blob/master/notebooks/diagnostic.ipynb), [code (c)](https://github.com/pachterlab/BLCSBGLKP_2020/blob/master/notebooks/lod_fda.ipynb), [code (d)](https://github.com/pachterlab/BLCSBGLKP_2020/blob/master/notebooks/viral_load.ipynb).

The FDA recommends that developers of diagnostic tests assess their method using a dilution series of three replicates per concentration with inactivated virus on actual patient specimens, and then confirm the final concentration with 20 replicates^14^. Based on this guidance, the FDA defines the limit of detection (LoD) as the lowest concentration at which 19/20 replicates are positive. Therefore, to assess the LoD from a standard curve, we performed 99 replicates of the training-testing and identified the titer at which the mean recall was equal to or above 0.95 (=19/20). The results (**Figure 2c**), can be automatically generated for any HMSA for which a standard curve has been generated. Moreover, by spiking in preset amounts of virus to make a standard curve alongside a group of samples being tested, our workflow makes possible dynamic calibration of the decision boundary for groups of samples being tested together. Finally, to test the ability of kallisto | bustools to estimate the amount of virus present, we fit a linear model to the virus counts and spike-in counts. We found a strong correlation between kallisto | bustools estimates and actual viral titer (**Figure 2d**). Estimation of viral load in the course of testing could help in determining time since infection^15^.

To validate our results we compared our approach to a complementary method which performed gene identification and sample index error correction using different algorithms. This alternative approach reverses the order of error correction of sample indices and assignment of biological reads to genes. First sample indices are identified and corrected using the Illumina utility bcl2fastq and then reads are clustered and the number of reads in each cluster are counted using starcode^16^. The bcl2fastq + starcode approach identifies slightly fewer aligned reads but otherwise produces results that are near identical to the kallisto | bustools results (**Figure 3**). However, in addition to mapping more reads, the kallisto | bustools workflow is faster, requires less memory, does not require a sample sheet, and is flexible to different barcoding assays.

**Figure 3:**
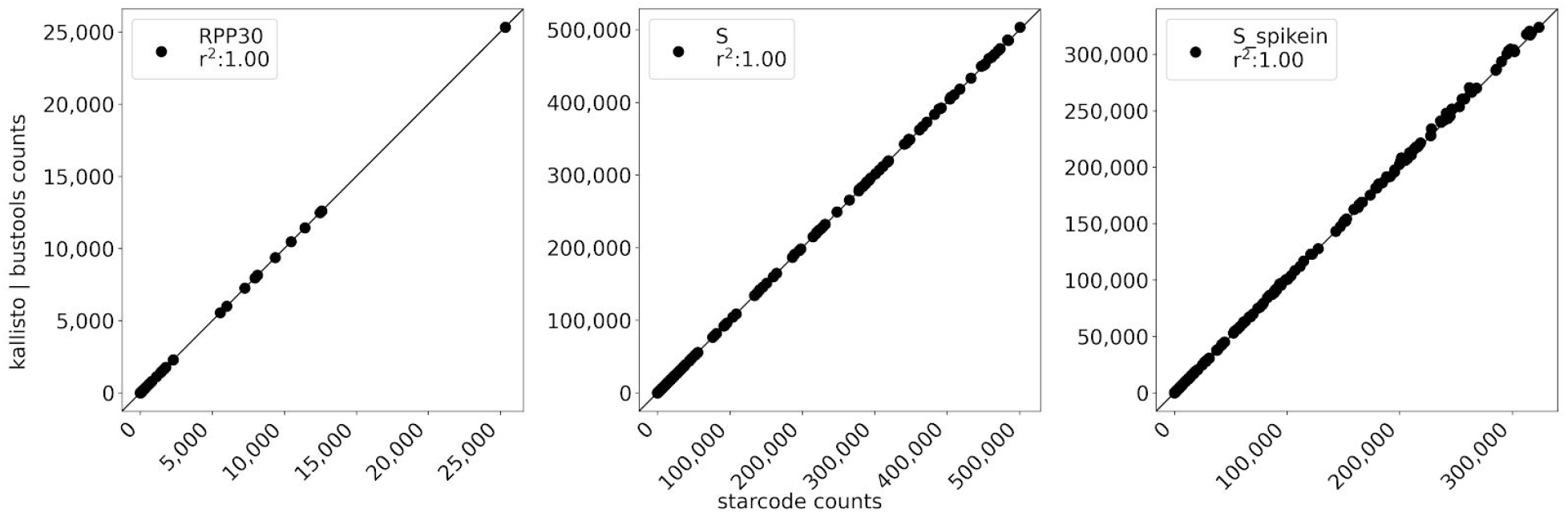
Orthogonal validation by read clustering. Scatter plots between kallisto | bustools workflow and starcode show near identical results on the genes targeted by the SwabSeq protocol a) RPP30, b) S, and c) S spike-in. Each point is a sample and the Pearson correlation is determined for the counts for a gene for all samples between kallisto | bustools and starcode. The code to reproduce this figure is here: [code](https://github.com/pachterlab/BLCSBGLKP_2020/blob/master/notebooks/kb_v_starcode.ipynb).

To illustrate the latter point, we extended kallisto to process three different diagnostic HMSAs: covE-seq, LAMP-seq, and TRB-seq data. To validate LAMP-seq and TRB-seq, we created two synthetic sets of sequencing reads which mimic the read structure of each assay. Starting with the count matrix from the SwabSeq assay and a set of 1,000 LAMP-seq sample indices^3^ we generated 12,062,027 single-end reads consisting of a gene target and a sample index (see Methods). Similarly, we used a set of 19,200 TRB-seq sample indices^4^ to generate paired-end reads, one for the sample index and one for the target gene. In both cases, bases in each read were randomly changed to another base with a probability of 0.005 to simulate Illumina sequencing errors. We processed these reads through kallisto | bustools and obtained near identical results to the kallisto | bustools results from the SwabSeq assay, thereby confirming the accuracy of the workflow for these assays (**Supplementary Figure 2** and **Supplementary Figure 3**). We also extended the kallisto | bustools workflow to process 2,437,573 reads produced with the covE-seq S5 protocol. The processing time was 8.17 seconds as compared to 20-22 hours with the covE-seq mBrave and BOLD cloud platforms^12^.

## Discussion

We have demonstrated a fast and accurate approach to processing highly multiplexed sequencing assays, and have validated an intuitive and interpretable method for obtaining diagnostic results from the data. Our workflow is easily extendable to assays which target different genes of interest and assays that have a different sample index structure. In addition, our approach is extendable to assays that incorporate unique molecular identifiers (UMIs) and that target regions of genetic variation, both of which are promising future directions for HMSAs.

The SwabSeq data shows that the data quality of HMSAs can be high and that accurate testing is technically feasible. The primary remaining challenge to widespread usage is, therefore, the organization of sample collection and associated logistics. While our work does not address the challenges of high-throughput sample collection, curation and handling of samples upstream of sequencing, our software does solve several post-sequencing logistics challenges. The low memory footprint of kallisto | bustools, specifically the requirement for less than 4Gb of RAM (**Supplementary Figure 7a**) enables essentially free compute in the cloud ^17^. Furthermore, the speed of the workflow allows for processing thousands of samples in minutes (**Supplementary Figure 7b**), which reduces overall testing time. The bustools software can also automatically identify sample indices without the need for a pre-configured Sample Sheet, thus facilitating quality control throughout the analysis. Finally, we have made the entire workflow easily usable on the cloud via Google Colaboratory which can be used to run the workflow for free via a browser window. This should facilitate collaborative optimization of analysis workflows, rapid deployment, and will simplify analysis logistics for large-scale testing.

While we have focused on SARS-CoV-2 testing in this manuscript, the methods we have developed are general and we expect that they should be applicable to future multiplexed diagnostic testing methods based on high-throughput sequencing.

## Data Availability

All the data, code and methods used to generate the results in this manuscript are open source freely available. All code to reproduce every figure and analysis for this manuscript is located here: https://github.com/pachterlab/BLCSBGLKP_2020. Each notebook can be run directly on Google Colab by pressing "Open in Colab" → "Runtime" → "Run all". The links to all FASTQ files can be found in Supplementary Table 1.
The SwabSeq protocol is described at https://www.notion.so/Octant-SwabSeq-Testing-9eb80e793d7e46348038aa80a5a901fd.
Software programs used are listed in Supplementary Table 2.

## Acknowledgments

We thank Páll Melsted for assistance with bustools.

## Author Contributions

ASB and LP developed the kallisto | bustools approach to processing and analyzing HMSA data. ASB modified kallisto and bustools to process SwabSeq, LAMP-seq, covE-seq, and TRB-seq data. ASB performed the analyses and collected results for the paper. NL developed the bcl2fastq + starcode processing approach with assistance from ARC, SWS, and JSB. JG assisted with technical aspects of the SwabSeq assay and in assessing the kallisto | bustools workflow results. LL created Figure 1 and explored the sample index structures of LAMP-seq, TRB-seq, and SwabSeq. SK, NLB, ARC, SWS and JSB developed SwabSeq. ASB and LP wrote the manuscript.

## Methods

### SwabSeq

The kallisto | bustools workflow was used to process an experiment with two 384-well plates. The wells included each of two different target genes, N and S, a varying amount of titered RNA from three different sources (Twist, ATCC RNA, and ATCC viral), a human gene control (RPP30), and two different lysates (HEK293, NP). Barcoded primers unique to each well, synthetic RNA spike-in controls that contained the same priming regions as the target RNA from SARS-CoV-2, primers for the target SARS-CoV-2 RNA, and a one-step RT-PCR mix were added to each well that RT-PCR was performed on. The wells were pooled and sequenced on an Illumina Nextseq.

### FASTQ files

Raw BCL files were converted into FASTQ files for the kallisto | bustools workflow using **bcl2fastq --create-fastq-for-index-reads** with read 1 corresponding to the Illumina i5 index, read 2 corresponding to the biological read, and index 1 corresponding to the Illumina i7 index.

### Alignment index

The genes targeted SwabSeq were a 108bp sequence of the SARS-CoV-2 S gene, a 6bp modification of the SARS-CoV-2 S gene (spike-in), a 72bp sequence of the SARS-CoV-2 N gene, a 6bp modification of the SARS-CoV-2 N gene (spike-in), and a 65bp sequence of the RPP30 gene (a housekeeping gene assumed to be present in all patient samples at a uniform abundance). The spike-in sequence differs from the original gene at the first 6bp. For each spike-in/viral gene, a 10bp window around the unique stretch of sequence was retained and the rest of the sequence removed since any 11mer that maps outside of the unique region will not be informative as to the origin of the read. A FASTA file of all hamming one distance variants of these target genes was made and indexed with **kallisto index -k 11** with a *k*-mer length of 11.

### Read alignment and BUS file processing

A BUS file is a columnar binary file where each row is a quadruplet of a sample index, UMI, set, and count that facilitates sample index error correction and amplicon quantification. Reads from the FASTQ files generated for the kallisto | bustools workflow were pseudoaligned using **kallisto bus -x SwabSeq** to generate a BUS file where all records in the UMI column are the same.

The BUS file was sorted with **bustools sort** which in addition to sorting the file lexicographically, counts and collapses the BUS records that are the same. Each half of the sample index in the BUS file was then corrected separately to a whitelist using **bustools correct -w whitelist.txt --split**. Correcting each half of the sample index independently is unique to SwabSeq which has a whitelist for the i5 and i7 primers, the combination of which makes the sample index. Each half of the sample index was corrected to at most Hamming distance one. The BUS file was sorted once more using **bustools sort** to count and collapse any additional BUS records that are the same.

The BUS file was converted to a text file and processed in Python to count the number of reads per sample that map uniquely to a specific target gene. This procedure yielded a sample by gene matrix where each entry corresponds to the number of reads for that sample that align to that gene. For the SwabSeq assay, this matrix consisted of 768 samples by 5 genes.

### Sample classification with logistic regression

Each sample was classified as + virus if it contained a non-zero amount of viral RNA. For each experiment, we split the data half into training and half into testing and used the viral, spike-in, and RPP30 counts as input. We learned the weights of a multivariate logistic regression model on the training data and used those weights to predict, for each sample, whether it contained virus. The logistic model used was

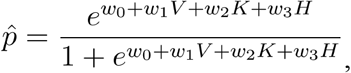

where V = log virus counts, K = log spike-in counts and H = log RPP30 counts.

### Viral load prediction with weighted linear regression

We performed a weighted linear regression on the viral and spike-in counts where the samples with known zero RNA titer were weighted by one over the number of unique titers. This was done to equalize the effect of each titer in the regression. For each experiment, we split the data into two halves: a training half and a testing half. The optimal coefficients for the linear model were learned from the training data, and the viral load was then predicted using those weights for the testing data. We performed this procedure on the log_2_(1 + counts). Given the weights *w_i_*, the known viral load *y_i_*, and the log of the counts for each training sample *X_ij_* plus one, the weighted linear regression model identified the vector of parameters *β* minimizing

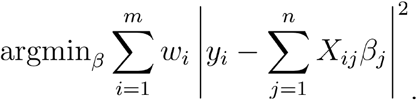

### Limit of detection

We iteratively removed the samples corresponding to increasing amounts of RNA titer, starting with the lowest titer, and performed 99 rounds split/test and logistic regression on the remaining samples. Each run reported the recall rate, i.e. the number of true positives divided by the sum of the number of true positives and false negatives. We defined the limit of detection (LoD) as the lowest RNA titer such that the mean of the recall for that titer is greater than or equal to 19/20 (0.95).

### Validation with a complementary bcl2fastq and starcode workflow

Demultiplexed FASTQ files for the orthogonal validation were generated using the sample sheet and **bcl2fastq --no-lane-splitting --sample-sheet SampleSheet.csv**. The default with bcl2fastq is error correction of 1 mismatch, so this step serves to error correct each index separately. The resultant demultiplexed FASTQ files corresponding to each sample were clustered using **starcode -d2 -t1 --sphere**. Sequence centroids were assigned to genes by exact matching, and the count for each gene was given by the number of reads up to Levenshtein distance 2 away from the centroid. A sample by gene matrix was then constructed from the counts.

### Generating of reads for testing LAMP-seq and TRB-seq

Reads were generated using the sample by genes matrix from the Plate 1, HEK293 lysate, N gene, Twist RNA SwabSeq experiment. This matrix consisted of 96 samples and 3 targeted genes. We obtained a list of 1000 sample indices from LAMP-Seq and 19,200 sample indices from TRB-Seq as well as their associated primer sets, N1, N2, and RPP30 for TRB-seq and B_B3 for LAMP-Seq. We generated a number of reads equal to the number of amplicon counts for each sample index and target gene pair, 12,062,027 reads in total. In addition, bases in each read were randomly mutated to another base with a probability of 0.005 to simulate Illumina sequencing errors. The read structure for TRB-seq is paired end reads with read 1 corresponding to a 15 basepair sample index and a 22 basepair constant region, and read 2 corresponding to the target gene. The read structure for LAMP-seq is single end reads where the first 20 basepairs correspond to the targeted gene, the next 22 basepairs correspond to the first forward inner primer, the subsequent 10 basepairs correspond to the sample index, and the last 19 basepairs correspond to the second forward inner primer.

### LAMP-seq and TRB-seq

Reads from each assay were processed with **kallisto bus -x LAMPSeq** and **kallisto bus -x TRBSeq** to generate a BUS file. The BUS file was sorted with **bustools sort**, the sample indices were corrected to Hamming distance 1 with **bustools correct** and the BUS file was sorted once more to sum duplicate records. The BUS file in text form was processed to generate the technology comparisons.

### covE-seq

We downloaded data in the form of FASTQ reads for the S5 protocol^12^, and processed the reads with **kallisto bus -x covEseq**. The processing time for all 2,437,573 reads was determined with the **time** command line utility.

### Data, protocol, and software availability

All the data, code and methods used to generate the results in this manuscript are open source freely available. All code to reproduce every figure and analysis for this manuscript is located here: https://github.com/pachterlab/BLCSBGLKP_2020. Each notebook can be run directly on Google Colab by pressing “Open in Colab” → “Runtime” → “Run all”. The links to all FASTQ files can be found in **Supplementary Table 1**.

The SwabSeq protocol is described at https://www.notion.so/Octant-SwabSeq-Testing-9eb80e793d7e46348038aa80a5a901fd.

Software programs used are listed in **Supplementary Table 2**.

